# Progressive Myocardial Flow Reserve Impairment Across Cardio-Kidney-Metabolic Syndrome Stages Is Associated with Cardiovascular Risk: Insights from Cardiac PET

**DOI:** 10.1101/2025.09.15.25335832

**Authors:** Filipe A. Moura, Alleh Nogueira, Joaquim Barreto, André Zimerman, Andrei C. Sposito, Robert Soufer, Aseem Vashist, Albert Sinusas, Edward Miller, Attila Feher

## Abstract

**Background:** Cardiovascular-Kidney-Metabolic (CKM) Syndrome is a progressive multisystem construct linked to elevated cardiovascular risk. Coronary microvascular dysfunction may reflect early CKM pathophysiology, but its prevalence and prognostic relevance across CKM stages are unclear. We hypothesized that myocardial flow reserve (MFR), measured by 82Rb PET/CT, declines with advancing CKM stage and provides prognostic value across the spectrum.

**Methods:** We retrospectively analyzed 5,502 patients who underwent rest-stress 82Rb PET myocardial perfusion imaging between 2016 and 2022. CKM stages 0–4 were defined using a structured algorithm integrating clinical, metabolic, and imaging features. Abnormal MFR was defined as <2.0. The primary outcome was a composite of all-cause death, myocardial infarction, stroke, or heart failure hospitalization. Associations between MFR, CKM stage, and outcomes were assessed using Cox models and Kaplan-Meier curves stratified by MFR and CKM stage.

**Results:** MFR declined stepwise with increasing CKM stage, from 2.65 ± 0.86 in Stage 0–1 to 2.08 ± 0.73 in Stage 4 (p < 0.001). Abnormal MFR prevalence rose from 21% to 50%. Over a median follow-up of 3.9 years (IQR 2.4–5.6), 1,527 composite events occurred. Both abnormal MFR (HR 2.23; 95% CI: 1.96–2.53) and higher CKM stage (Stage 4 vs. Stage 0–1: HR 2.99; 95% CI: 1.71–5.21) independently predicted events. Abnormal MFR further stratified risk within each CKM stage, with the strongest association in Stage 0–1 (HR 7.26; 95% CI: 2.36–22.4; p-interaction < 0.05).

**Conclusions:** MFR is progressively lower across CKM syndrome stages. Abnormal MFR is independently associated with cardiovascular events and provides incremental prognostic value within each CKM stage—most notably in early-stage disease—supporting its potential role in improving risk stratification across the CKM continuum.

## Introduction

The rising global burden of cardiovascular disease (CVD), chronic kidney disease, and metabolic disorders such as type 2 diabetes and obesity has prompted a shift toward integrative frameworks that reflect the shared biological and clinical pathways underlying these conditions. One such framework is the Cardiovascular-Kidney-Metabolic (CKM) Syndrome. This staged classification system progresses from early subclinical risk factors to established end-organ disease, aiming to facilitate earlier detection, holistic risk stratification, and coordinated care^1^. Almost 90% of US adults met the criteria for earlier stages of CKM, and 15% met the requirements for more advanced stages^2^.

CKM syndrome is rooted in shared pathophysiological mechanisms, most notably chronic inflammation, insulin resistance, endothelial dysfunction, and adverse vascular remodeling^1,3,4^. A key manifestation of these processes is impaired coronary microvascular function – reflected partly in impaired myocardial blood flow (MBF) and myocardial flow reserve (MFR) –, which has emerged as a critical, yet relatively underrecognized contributor to CVD risk, including individuals without obstructive coronary artery disease, hypertension, CKD, and obesity^5–8^. While existing data suggest that a greater number of cardiometabolic risk factors and increased atherosclerotic burden are associated with impaired coronary microvascular function^9^, the association of these markers of overall coronary vascular health (MBF and MFR) with CKM Syndrome, and their subsequent prognostic relevance within the CKM construct, remains unknown.

In this context, we aimed to (1) assess the burden of impaired MFR across CKM syndrome stages and (2) determine its prognostic value within CKM stages in a large and diverse cohort of patients referred for rest-stress PET myocardial perfusion imaging. We hypothesized a higher prevalence of abnormal MFR with increasing CKM stage and that abnormal MFR would be associated with excess cardiovascular risk across the CKM spectrum.

## Methods

### Study Population

We conducted a retrospective cohort study from a single-site clinical registry of patients referred for a nuclear stress test with positron emission tomography with contrast tomography for attenuation correction (PET/CT) at Yale New Haven Hospital, Connecticut, between 2016 and 2022. We analyzed 5,749 patients who underwent stress testing with regadenoson PET/CT with ^82^Rubidium (^82^Rb). For patients with multiple PET studies, only the earliest exam was retained to ensure independent observations. Patients with prior heart transplantation were excluded. The study was approved by the institutional review board (HIC# 2000021621) and conducted in accordance with the ethical standards of the responsible institution and the principles of the Declaration of Helsinki.

### PET Imaging Protocol

Patients were instructed to refrain from taking caffeine or methylxanthine-containing substances for at least twelve hours before PET imaging. Dynamic rest-stress ^82^Rb PET myocardial perfusion imaging was performed using either a hybrid PET/CT scanner with a 16-slice CT component (Discovery ST, GE Healthcare) or a 64-slice CT component (Discovery 690, GE Healthcare), as previously described^10–12^. Briefly, rest imaging was conducted following intravenous administration of ^82^Rb. Pharmacologic stress by vasodilator hyperemia was then induced with regadenoson 0.4 mg IV bolus. At peak stress, ^82^Rb was administered via peripheral IV, and dynamic stress images were acquired. Low-dose CT imaging was performed for attenuation correction of the PET data.

### PET/CT Data Analysis

PET images were reconstructed with attenuation correction using system software to generate dynamic image series, which were then reprocessed and analyzed using Invia Corridor 4DM v2017 (Ann Arbor, MI). The summed stress score (SSS), summed rest score, and summed difference score were also calculated. Abnormal perfusion was considered when SSS was greater than 2. Rest LVEF was obtained from gated myocardial perfusion images.

Quantitative global MBF (mL/min/g) was extracted at both rest and stress from dynamic images that were fitted into a validated two-compartment kinetic model as previously described.^13^ MFR was calculated as the ratio of stress to rest MBF. Abnormal stress MBF and MFR were defined as values <1.8 mL/min/g and <2.0, respectively. Coronary artery calcification (CAC) and aorta calcification were qualitatively graded by experienced nuclear readers on the day of the study on a scale ranging from none, mild, moderate, to severe for each coronary artery (left main, left anterior descending, circumflex, and right) and thoracic aorta segment (ascending or descending). Coronary or aorta calcification was considered present when any single coronary or thoracic aorta segment was noted to have at least mild calcification on the attenuation CT.

### Variable Harmonization and Clinical Feature Extraction and Definitions

Demographic variables, clinical comorbidities, outpatient medications, and cardiovascular risk factors – including hypertension, hyperlipidemia, diabetes, prediabetes, metabolic syndrome, chronic kidney disease (CKD), end-stage kidney disease (ESKD), atrial fibrillation, and heart failure (HF) – were defined using both structured variables and free clinical note text from the registry report forms. Text fields were parsed using regular expressions to identify relevant diagnostic terms (Supplement). Similarly, the presence of atherosclerotic cardiovascular disease (ASCVD) was defined based on coded fields and text references to myocardial infarction (MI), stroke, peripheral artery disease, and coronary revascularization. Subclinical cardiovascular disease was defined as either subclinical ASCVD – i.e., (1) abnormal perfusion or (2) coronary or aortic calcification on PET/CT imaging in the absence of known ASCVD or symptoms – or subclinical heart failure – left ventricular ejection fraction less than 40% on resting gated PET/CT in the absence of a documented history of HF. Aortic calcification was considered an equivalent of subclinical ASCVD given known similar associations with excess cardiovascular and total mortality when compared to CAC^14^. Excess adiposity was defined as a BMI greater than or equal to 25 kg/m^2^ for non-Asian individuals and greater than or equal to 23 kg/m^2^ for Asian individuals.^15^

### CKM Syndrome Classification

Patients were categorized into mutually exclusive stages of CKM syndrome using a hierarchical, rule-based algorithm that integrated clinical disease status, metabolic risk factors, and imaging-derived markers of subclinical disease:

● Stage 4: Defined by the presence of established clinical cardiovascular disease – history of HF, ASCVD (MI, stroke, PAD, or prior CABG or PCI), or atrial fibrillation – along with at least excess adiposity, hypertension, hyperlipidemia, diabetes, metabolic syndrome, or CKD).
● Stage 3: Defined by the absence of manifest cardiovascular disease, but the presence of either subclinical atherosclerotic cardiovascular disease (abnormal perfusion or any presence of coronary or aortic calcification without clinical ASCVD), subclinical heart failure (resting left ventricular function less than 40% on gated PET/CT images, without a history of HF), or ESKD (dialysis, ESKD terms, or history of kidney transplant), in addition to at least one of the following: excess adiposity, hypertension, hyperlipidemia, diabetes, or metabolic syndrome.
● Stage 2: Defined by the absence of both manifest and subclinical cardiovascular disease, and the presence of either CKD (excluding ESKD), hypertension, hyperlipidemia, diabetes, or metabolic syndrome.
● Stage 1: Defined by the absence of both manifest and subclinical cardiovascular disease, CKD, hypertension, hyperlipidemia, diabetes, metabolic syndrome, and the presence of either excess adiposity or prediabetes.
● Stage 0: Defined by the absence of all the above – absence of manifest and subclinical cardiovascular disease, CKD, excess adiposity, prediabetes, or other cardiometabolic risk factors.

### Clinical Outcomes and Follow-Up

Patient follow-up data were obtained from outpatient visits, hospital admissions, and the electronic medical record. A composite cardiovascular outcome was defined as the first occurrence of death, MI, stroke, or HF hospitalization. Follow-up time was calculated as the time from PET imaging to the event or the last available clinical contact.

### Statistical Analyses

Descriptive statistics were calculated to summarize baseline characteristics, stratified by CKM syndrome stage. Continuous variables were expressed as mean ± standard deviation (SD) for normally distributed data or as median with interquartile range (IQR) for non-normally distributed data. Categorical variables were summarized as counts and percentages.

CKM stages 0 and 1 were combined, given the group size and comparable cardiovascular risk between stages. Comparisons across CKM syndrome stages were performed with one-way analysis of variance (ANOVA) for continuous variables and chi-square or Fisher’s exact test for categorical variables.

Global rest and stress MBF and MFR were analyzed both as continuous variables and as binary indicators based on predefined thresholds: MFR <2.0 and stress MBF <1.8 mL/g/min were considered abnormal. We generated a derived categorical variable reflecting combinations of MFR and stress MBF status to explore discordant or concordant impairments across CKM syndrome stages. Stratified summary statistics were also computed after excluding patients with abnormal perfusion defects on PET imaging to isolate the prognostic relevance of flow metrics independent of meaningful ischemia.

Event rates of the composite endpoint and each of the sub-components were compared between groups defined by abnormal vs. normal MFR, and within each CKM stage subgroups. Kaplan-Meier survival curves for the composite endpoint were generated, and comparisons between groups were conducted using the log-rank test. Univariable and multivariable Cox proportional hazards regression models were constructed to evaluate the association between abnormal MFR and clinical outcomes. Models were adjusted for age and sex, with hazard ratios (HRs) and 95% confidence intervals (CIs) reported. Cox proportional hazards regression models were conducted for the overall population, for patients with normal perfusion, and stratified by CKM stage. Interaction terms were used to assess whether the association between MFR and outcomes differed across CKM stages.

Additionally, we modeled the CKM stage as both a categorical and an ordinal variable to test for trends in event risk across stages. Combined models including CKM stage and MFR (categorical or continuous) were evaluated to assess additive prognostic values. Subgroup-specific adjusted models were also estimated to derive stage-specific hazard ratios for abnormal MFR, both in the full cohort and restricted to patients with normal perfusion, to support stratified analyses. To assess the incremental prognostic value of MFR beyond CKM stage, we compared nested logistic regression models using the area under the receiver operating characteristic curve (AUC), integrated discrimination improvement (IDI), and continuous net reclassification improvement (NRI), with 1,000 bootstrap iterations used to estimate 95% confidence intervals. A two-sided p-value <0.05 was considered statistically significant. All statistical analyses were conducted using R version 4.4.2

## Results

### Baseline Characteristics

A total of 5,502 of the 5,749 patients who underwent ^82^Rb PET/CT myocardial perfusion imaging were categorized by CKM Syndrome stage and included in the analytic cohort. Overall, the mean age was 64 ± 12 years, with 52% being women, and 44% being non-White (**Table 1**). The majority had CKM risk factors, and 42% and 18% had clinical and subclinical ASCVD, respectively. The prevalence of individuals in each stage of CKM syndrome was as follows: Stage 0 or 1 (n=226; 4.1%), Stage 2 (n=827; 15.0%), Stage 3 (n=1,434; 26.1%), and Stage 4 (n=3,015; 54.8%). Higher CKM stage was associated with older age, greater prevalence of male sex, and a greater burden of cardiometabolic comorbidities (as per CKM stage definition). As per definition, diabetes, hypertension, and hyperlipidemia were nearly absent in CKM Stage 0 or 1 but present in 42–83% of patients in Stage 2 through 4.

**Table 1.**
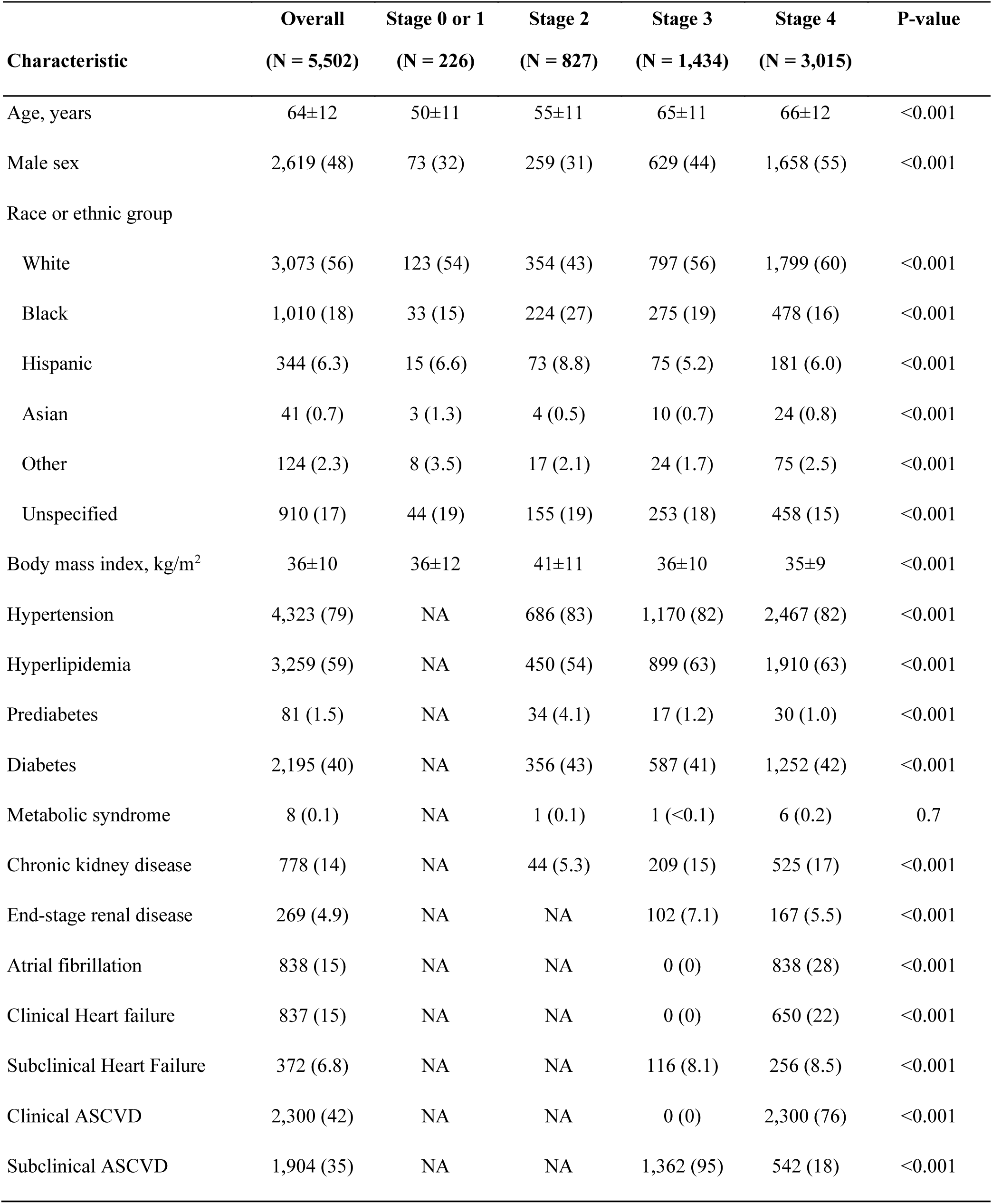
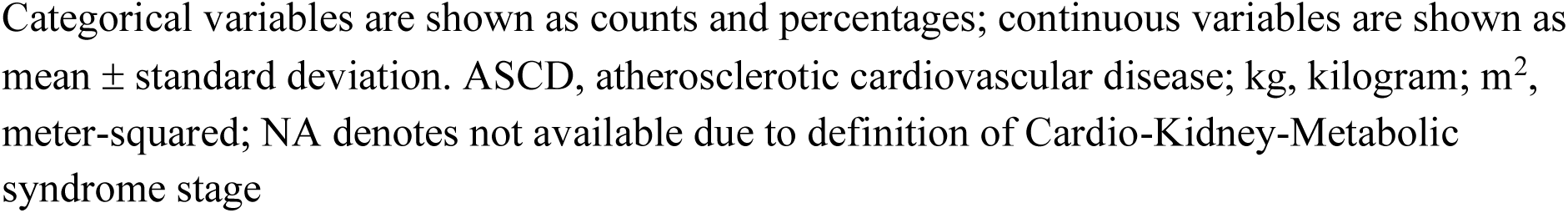
Baseline Patient Characteristics Across Stages of Cardio-Kidney-Metabolic Syndrome

### Myocardial Blood Flow and Flow Reserve Across CKM Stages

MFR and stress MBF demonstrated a stepwise decline with advancing CKM syndrome stage (**Table 2**). Mean MFR decreased from 2.65 ± 0.86 in CKM Stage 0 or 1 to 2.08 ± 0.73 in Stage 4 (p < 0.001). The proportion of patients with abnormal MFR, defined as <2.0, also rose across CKM stages—from 21% in Stage 0 or 1 to 50% in Stage 4 (**Table 2**). Similarly, the prevalence of abnormal stress MBF (<1.8 mL/min/g) rose from 14% in Stage 0 or 1 to 47% in Stage 4 (p < 0.001). Patients with more advanced CKM stages more frequently had concordant abnormal MFR and stress MBF (5.9% in Stage 0 or 1 vs. 35% in Stage 4). These significant trends persisted even among patients with visually normal perfusion (**Supplementary Table 1**).

**Table 2.** Myocardial Blood Flow and Myocardial Flow Reserve According to CKM Syndrome Stage.

| Characteristic | Overall<br>(N = 5,502) | Stage 0 or 1<br>(N = 226) | Stage 2<br>(N = 827) | Stage 3<br>(N = 1,434) | Stage 4<br>(N = 3,015) | P-value |
| --- | --- | --- | --- | --- | --- | --- |
| MFR | 2.22±0.74 | 2.65±0.86 | 2.53±0.69 | 2.25±0.70 | 2.08±0.73 | <0.001 |
| Abnormal MFR | 2,212 (41) | 47 (21) | 168 (21) | 540 (39) | 1,457 (50) | <0.001 |
| Stress MBF, mL/min/g | 2.21±0.90 | 2.71±0.95 | 2.73±0.99 | 2.33±0.82 | 1.98±0.82 | <0.001 |
| Abnormal stress MBF | 1,868 (35) | 30 (14) | 115 (14) | 361 (26) | 1,362 (47) | <0.001 |
| Rest MBF, mL/min/g | 1.04±0.41 | 1.09±0.46 | 1.13±0.44 | 1.09±0.42 | 0.99±0.38 | <0.001 |
| Combined MFR and Stress MBF |  |  |  |  |  | <0.001 |
| Normal MFR and stress MBF | 2,572 (48) | 158 (71) | 586 (72) | 726 (52) | 1,102 (38) |  |
| Normal MFR and abnormal stress MBF | 550 (10) | 17 (7.7) | 58 (7.1) | 123 (8.9) | 352 (12) |  |
| Abnormal MFR and normal stress MBF | 886 (17) | 34 (15) | 111 (14) | 297 (21) | 444 (15) |  |
| Abnormal MFR and stress MBF | 1,317 (25) | 13 (5.9) | 57 (7.0) | 238 (17) | 1,009 (35) |  |

CKM, Cardio-Kidney-Metabolic; MBF, myocardial blood flow; MFR, myocardial flow reserve.

### Cardiovascular Outcomes and Risk Stratification by MFR and CKM Stage

Overall, there were a total of 1527 composite cardiac outcomes (829 deaths, 392 MIs, 214 strokes, and 821 HF admissions) over a median follow-up of 3.9 years (IQR 2.4–5.6).

Patients with abnormal MFR experienced 2.5-fold higher event rates of the primary composite outcome (death, myocardial infarction, stroke, or heart failure) and significantly greater event rates for each of its subcomponents (**Table 3**). In the overall cohort, the association between abnormal MFR and outcomes remained significant after adjustment for CKM stage (**Table 3**: HR 2.23; 95% CI: 1.96–2.53) and persisted when MFR was analyzed as a continuous variable (**Supplementary Table 2:** per MFR unit decrease HR 2.17; 95% CI, 1.96–2.44). In patients with normal perfusion, there were also greater hazards for the primary endpoint even after adjusting for age, sex, and CKM stage (**Supplementary Table 3**: HR 2.05; 95% CI 1.73–2.42 for abnormal MFR and per MFR unit decrease HR 1.89; 95% CI, 1.64–2.17).

**Table 3.** Cardiovascular Outcomes According to MFR Status.

| Group | Normal MFR |  |  | Abnormal MFR |  |  | Hazard Ratio | P-value |
| --- | --- | --- | --- | --- | --- | --- | --- | --- |
|  | Events/No of patients (%) | Patient-years | Events per 100 patient-years | Events/No of patients (%) | Patient-years | Events per 100 patient-years |  |  |
| Overall |  |  |  |  |  |  |  |  |
| Stage 0 or 1 | 5/173 (2.9) | 629.3 | 0.8 | 8/40 (20.0) | 130.3 | 6.1 | 7.26 (2.36-22.4) | <0.001 |
| Stage 2 | 50/636 (7.9) | 2545.4 | 2.0 | 22/153 (14.4) | 566.5 | 3.9 | 1.91 (1.15-3.16) | 0.012 |
| Stage 3 | 103/807 (12.8) | 3018.6 | 3.4 | 109/477 (22.9) | 1625.4 | 6.7 | 1.92 (1.46-2.51) | <0.001 |
| Stage 4 | 261/1385 (18.8) | 5002.0 | 5.2 | 486/1207 (40.3) | 3528.7 | 5.2 | 2.33 (2.00-2.72) |  |
| Normal perfusion |  |  |  |  |  |  |  |  |
| Stage 0 or 1 | 5/173 (2.9) | 629.3 | 0.8 | 7/39 (17.9) | 130.0 | 5.4 | 6.64 (2.10-21.0) | <0.001 |
| Stage 2 | 50/635 (7.9) | 2539.0 | 2.0 | 22/153 (14.4) | 566.5 | 3.9 | 1.90 (1.15-3.15) | 0.012 |
| Stage 3 | 79/627 (12.6) | 2396.2 | 3.3 | 73/324 (22.5) | 1148.0 | 6.4 | 1.92 (1.40-2.65) | <0.001 |
| Stage 4 | 166/965 (17.2) | 3581.1 | 4.6 | 184/515 (35.7) | 1685.6 | 10.9 | 2.06 (1.66-2.57) | <0.001 |

Abnormal myocardial flow reserve (MFR) was defined as less than 2.0. The primary outcome was a composite of death from any cause, myocardial infarction, stroke, or heart failure.

### Interaction Between MFR and CKM Stage for Cardiovascular Risk

Advancing CKM stage was associated with a graded increase in risk for the composite outcome (adjusted HR for Stage 4 vs. Stage 0 or 1: 2.99; 95% CI: 1.71–5.21; **Supplementary Table 2**). Kaplan-Meier curves demonstrated increasing cumulative incidence of the primary outcome with higher CKM stage (log-rank p <0.001) (**Figure 1**). Trend analyses confirmed a significant linear association across CKM stages, even after adjusting for age, sex, and MFR (p for trend <0.001). Patients with concurrent abnormalities in both MFR and stress MBF had nominally highest event rates across all CKM stages 0 or 1, 3, and 4 (**Supplementary** Figure 1). For example, in Stage 4, the annualized event rate for the primary outcome was 15.4% of patients with both abnormal MFR and stress MBF compared with 4.8% in those with normal values for both. The additive prognostic value of dual impairment was also evident in lower-risk CKM stages.

**Figure 1.**
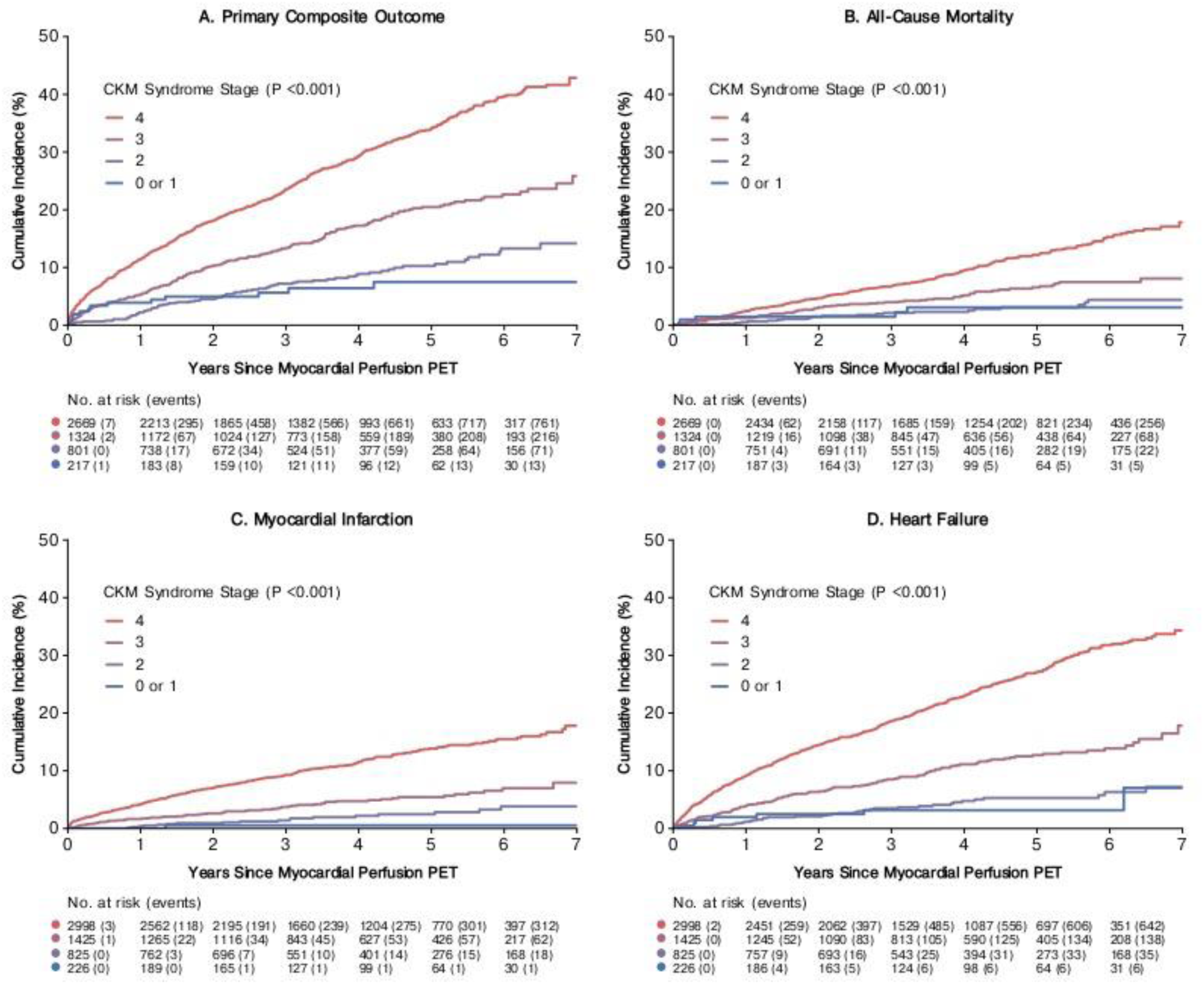
Cumulative incidence of (A) composite, (B) death, (C) myocardial infarction, and (D) heart failure, according to CKM Syndrome Stage.

The prognostic impact of abnormal MFR varied by CKM stage (**Figure 2A**). Adjusted interaction terms between abnormal MFR and CKM stages were all statistically significant (p < 0.045), indicating a relative attenuation of risk in Stages 2 through 4 compared with the reference group (Stage 0 or 1). Among individuals in Stage 0 or 1, abnormal MFR conferred a greater than 7-fold higher risk of the primary outcome (HR 7.26; 95% CI: 2.36–22.4), with an absolute risk comparable to that of patients in Stage 4 CKM with normal MFR (6.1 vs. 5.2 events per 100 patient-years). This association was progressively attenuated in more advanced CKM stages, with interaction terms suggesting a significantly lower relative hazard in Stages 2 through 4 (**Supplementary** Figure 1). Although attenuated, elevated risk persisted in Stage 2 (HR 1.91; 95% CI: 1.15–3.16), Stage 3 (HR 1.92; 95% CI: 1.46–2.51), and Stage 4 (HR 2.33; 95% CI: 2.00–2.72). Similar findings were observed in sensitivity analyses limited to patients with normal perfusion (**Figure 2B**). Kaplan-Meier curves demonstrated significantly different and increasing cumulative incidence of the primary outcome with abnormal MFR within each CKM stage (**Figure 3**). Lastly, to evaluate the incremental prognostic value of MFR, we compared nested models using discrimination and reclassification metrics. Adding MFR to CKM stage improved discrimination from AUC = 0.63 to AUC = 0.70 (p < 2×10⁻¹⁶; **Supplementary** Figure 2), with an integrated discrimination improvement (IDI) of 0.034 (95% CI: 0.029–0.040; p < 0.001). Risk reclassification was also enhanced, with a continuous net reclassification improvement (NRI) of 0.32 (95% CI: 0.26–0.35; p < 0.001).

**Figure 2.**
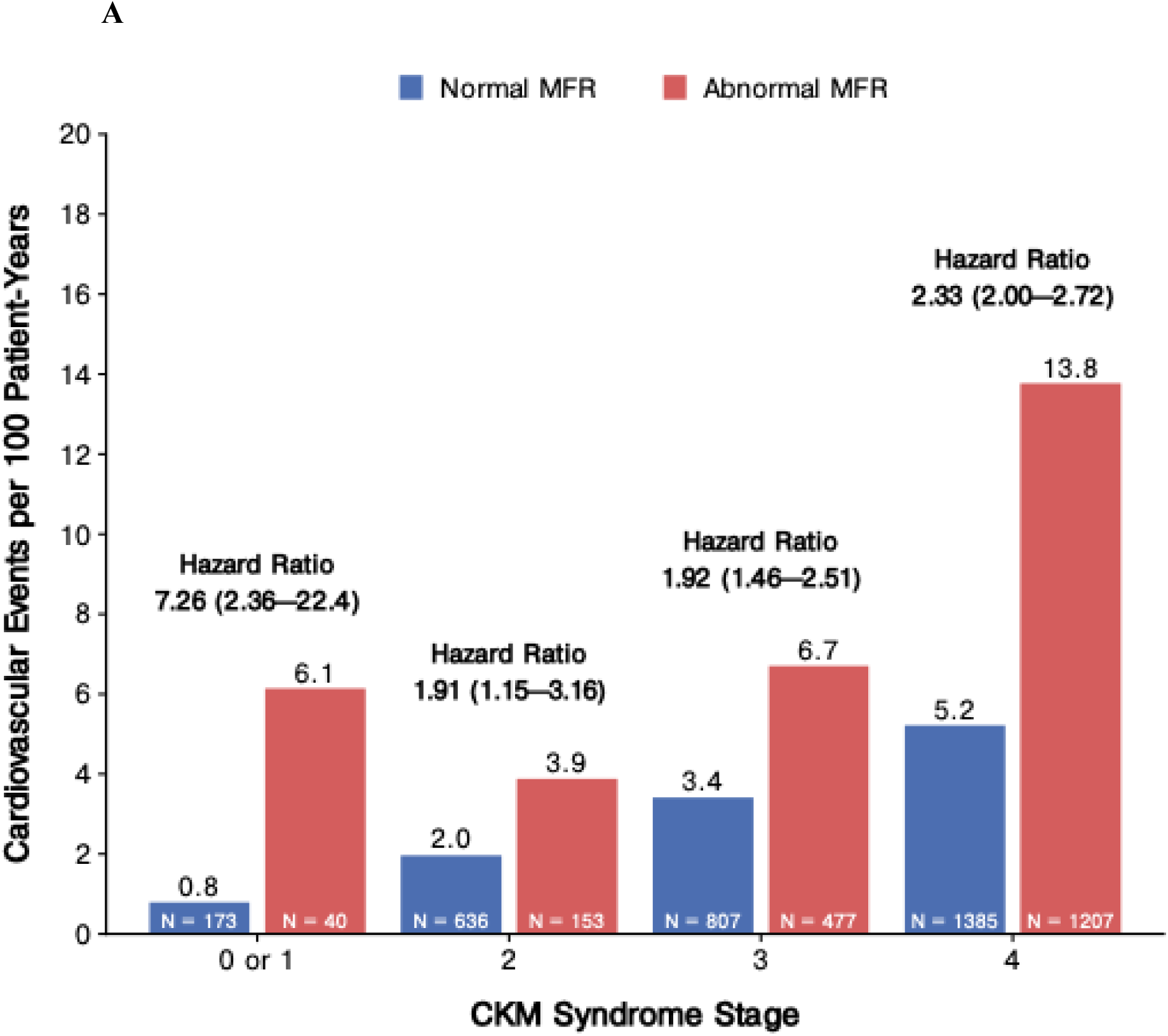
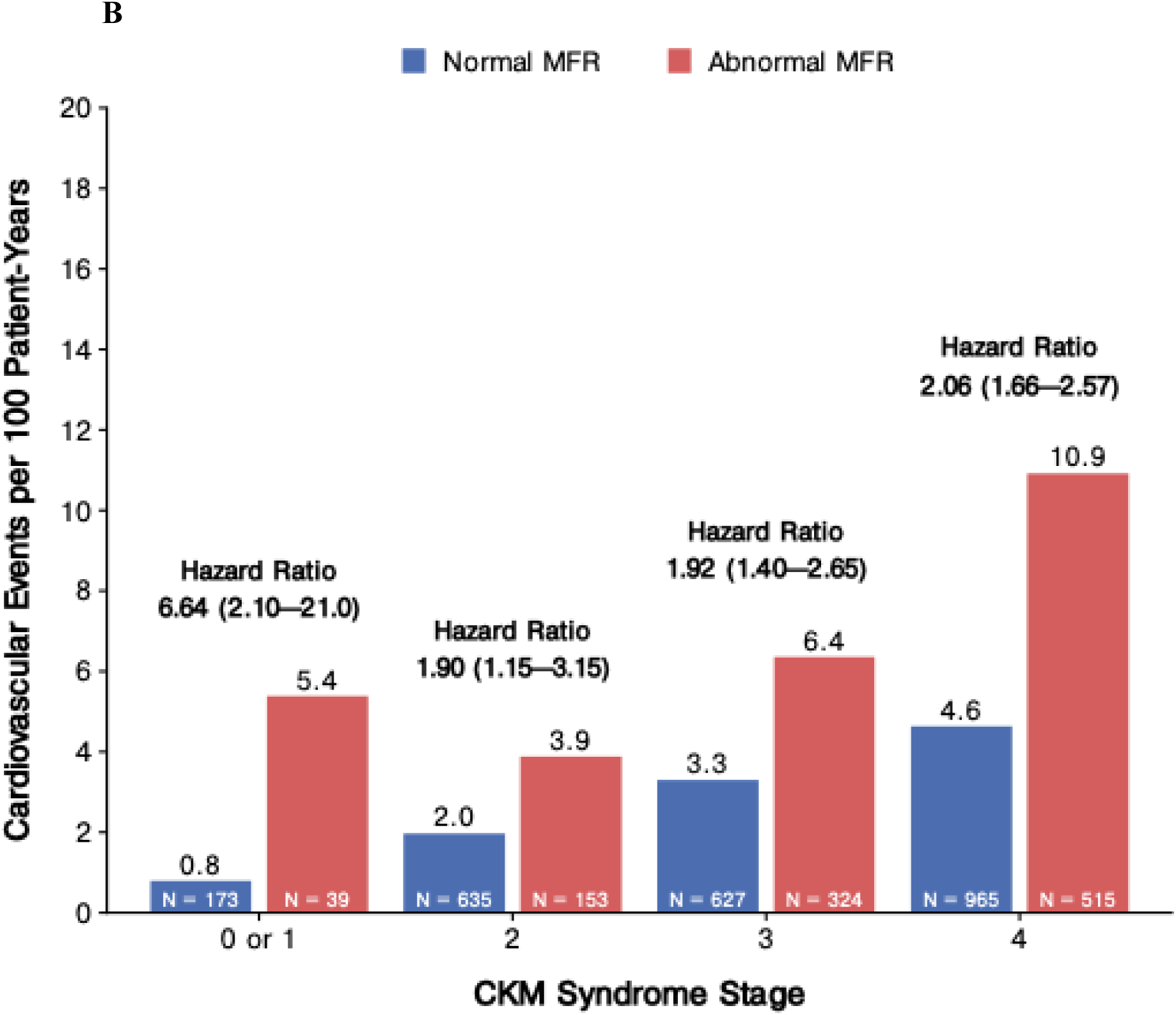
Annualized event rates of composite cardiovascular outcomes across the stages of CKM Syndrome according to MFR. Incidence and risk (HR 95% confidence interval) of composite cardiovascular outcomes (death, myocardial infarction, stroke, and heart failure) in patients with abnormal MFR compared to those with normal MFR according to CKM syndrome stages in (A) overall and (B) normal perfusion cohort.

**Figure 3.**
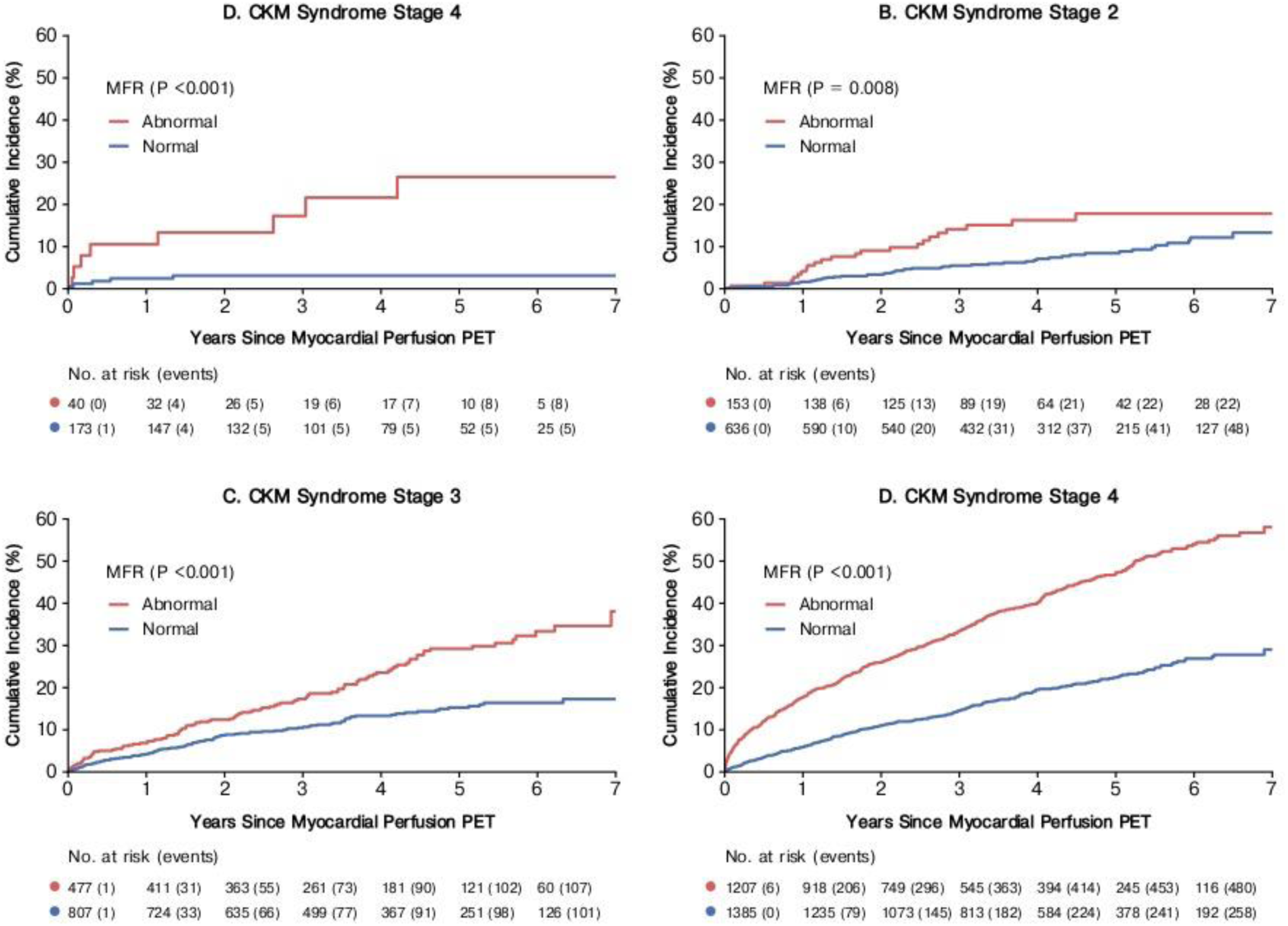
Cumulative incidence of composite cardiovascular outcome in patients with normal and abnormal MFR according to each stage of CKM Syndrome.

## Discussion

In this large cohort study of patients undergoing clinically indicated PET/CT myocardial perfusion imaging, we provide the first comprehensive assessment of MFR, a representative of coronary epicardial obstruction and microvascular function, across the spectrum of the CKM syndrome. Our findings demonstrate several key insights. First, we observed a stepwise decline in both stress MBF and MFR with advancing CKM stage, with a corresponding increase in the prevalence of impaired stress MBF and MFR, including among patients without clear perfusion defects. Second, we confirmed that a more advanced CKM stage was independently associated with adverse cardiovascular outcomes and that both CKM stage and MFR provided additive prognostic information. Third, we found that the prognostic value of abnormal MFR was present across all CKM stages but was most pronounced in the earliest stages.

Coronary epicardial obstruction and microvascular dysfunction, as measured by MFR, are well-established features of cardiometabolic disease and CKD. In hypertension, microvascular impairment arises from endothelial dysfunction, oxidative stress, and structural remodeling of the arteriolar network^16^, and is associated with elevated systolic blood pressure, increased left ventricular mass, and a heightened risk of heart failure and adverse cardiac events^17–19^. Obesity contributes to microvascular dysfunction through chronic low-grade inflammation, adipocytokine-mediated endothelial injury, and microvascular remodeling^20,21^. MFR decreases linearly with increasing BMI, and impaired MFR is a stronger predictor of adverse cardiovascular outcomes than BMI itself^22^. Similarly, diabetes induces early and progressive microvascular dysfunction via hyperglycemia-induced endothelial injury, increased oxidative stress, and microvascular rarefaction^23^, with consistent reductions in MFR observed across multiple studies^24–26^. In CKD, impaired MFR is evident from early CKD stages, reflecting microvascular dysfunction, and is independently associated with major adverse cardiovascular events regardless of kidney function^27,28^.

Taken together, this body of evidence aligns with our study’s findings: within the CKM construct—which spans from early cardio-kidney-metabolic risk to manifest cardiovascular disease such as obstructive coronary artery disease (CAD) or heart failure— we observed a progressive decline in MFR and MBF across advancing stages. This trend was evident both in patients with abnormal perfusion, likely reflecting obstructive CAD, and in those with normal perfusion, highlighting a predominant contribution of escalating coronary microvascular dysfunction in parallel with worsening multisystem risk. Furthermore, the independent prognostic value of MFR within each CKM stage underscores its relevance as a unifying marker of cardiovascular risk across this spectrum. Future research should explore whether integrating MFR into CKM-based risk stratification can improve clinical decision-making and guide preventive or therapeutic interventions.

Notably, individuals with Stage 0 or 1 CKM and abnormal MFR had a similar or even higher risk of adverse cardiovascular events than those in later CKM stages with preserved MFR, suggesting that MFR can unmask elevated risk in patients otherwise classified as low risk. While these findings support the relevance of coronary microvascular dysfunction within the CKM construct, they do not imply causality. Instead, they may reflect complex, bidirectional relationships among metabolic dysfunction, endothelial injury, and impaired coronary vasodilation. For example, insulin resistance and chronic inflammation in early metabolic syndrome may contribute to reduced hyperemic response. The presence of microvascular dysfunction in early CKM stages may also indicate an alternate, subclinical disease axis not fully captured by CKM staging but nonetheless associated with increased risk. These hypotheses warrant further longitudinal and mechanistic studies.

This study had some limitations. First, the analysis was retrospective and conducted at a single center, which may limit generalizability and introduce the potential for selection bias. For instance, the cohort was composed of patients referred for clinically indicated PET myocardial perfusion imaging, most commonly for symptoms or cardiovascular risk stratification. As such, our study population represents a higher-risk population with limited representation of individuals in earlier CKM stages. Only 4.1% of our study population were classified as stage 0 or 1, which may have led to overestimation of the risk of abnormal MFR in this subgroup. Furthermore, the utility of MFR as a prognostic tool in asymptomatic and early-stage CKM—who may not undergo stress testing in routine practice—remains to be determined. Second, the classification of CKM syndrome stages was constrained by the available data. Specifically, we lacked laboratory values such as hemoglobin A1c, lipids, and renal injury and function assessments, which limited our ability to identify undiagnosed cardio-kidney-metabolic conditions like diabetes, metabolic syndrome, CKD, and to use this data for further cardiovascular risk assessment with risk scores^4^. Additionally, body mass index was used as a surrogate for excess adiposity; more refined anthropometric measures (e.g., waist circumference, visceral fat assessment) were not available and may have provided more accurate CKM classification. Finally, while PET-derived MFR and MBF provide robust physiologic insights—even in patients with visually normal perfusion—they may still be influenced by diffuse, non-obstructive coronary artery disease. As such, a more definitive assessment of coronary microvascular dysfunction would ideally include invasive physiologic testing.

In conclusion, in this large, retrospective cohort of patients undergoing rest-stress PET myocardial perfusion imaging, we demonstrate that MFR and stress MBF decline progressively with advancing CKM syndrome stages, aligning with the multisystem pathobiology underlying the CKM construct. In addition, our findings highlight that both CKM stage and MFR are independently associated with adverse cardiovascular outcomes, and that MFR adds prognostic value even within individual CKM stages, helping identify patients at particularly high cardiovascular risk.

## Data Availability

We encourage parties interested in collaboration and data sharing to contact the corresponding author directly for further discussions.

## ACKNOWLEDGEMENTS

Funding and Assistance:

This study had no source of funding.

## Author Contributions and Guarantor Statement

F.A.M. conceptualized and designed the study. A.N. and F.A.M. analyzed the data. All authors were involved in data interpretation. F.A.M. drafted the manuscript, and all authors revised the manuscript. All authors approved the final submitted version and agreed to be accountable for the report. Dr. Filipe A. Moura and Attila Feher are the guarantors of this work and, as such, had full access to the data in the study and take responsibility for the integrity of the data and the accuracy of the data analysis.

